# Incidence, outcome, and dynamics of out-of-hospital cardiac arrest in the city of Vienna between 2019 and 2023

**DOI:** 10.1101/2025.04.12.25325712

**Authors:** Mario Krammel, Andrea Kornfehl, Daniel Grassmann, Thomas Hamp, Thomas Grubmiller, Alexander Nürnberger, Hans Domanovits, Patrick Aigner, Michael Girsa, Patrick Glaninger, Andreas Zajicek, Patrick Sulzgruber, Michael Holzer, Sebastian Schnaubelt

## Abstract

**Background:** In coherent regions, key figures and trends of out-of-hospital cardiac arrest (OHCA) are vital to improve favourable outcomes. Since the last cardiopulmonary resuscitation (CPR) guideline update, comprehensive OHCA data of the metropolitan area of Vienna, Austria, have been scarce.

**Methods:** This retrospective study analyzed adult non-traumatic OHCA cases in Vienna between 01/2019 and 12/2023. It assessed emergency medical service records and clinical patient data, and reports incidences, ROSC rates, survival to hospital discharge, and neurological outcome. Logistic regression assessed associations between outcomes and predictors, while Poisson regression examined incidence changes before, during, and after COVID-19 lockdowns.

**Results:** During the observation period, the Emergency Medical Service Vienna started cardiopulmonary resuscitation (CPR) in a total of 7433 patients (77.1/100,000 population per year). Sustained return of spontaneous circulation (ROSC) was observed in 24.8%, survival to hospital discharge in 9.3%, and a Cerebral Performance Category Score (CPC) of 1 or 2 in 6.8%. Compared with previous literature, outcomes remained stable. However, in patients with witnessed cardiac arrest of suspected cardiac aetiology and an initial shockable rhythm, rates amounted up to 39% for hospital discharge and 29.6% for CPC 1 or 2. Similarly, patients with CPC 1 or 2 before CPR had better outcomes than the overall cohort. During COVID-19, there was a decline in all outcome parameters.

**Conclusions:** Survival after OHCA in Vienna seems stable, but significant improvements in outcome parameters are seen in a ‘high outcome potential cohort’ over the last 15 years. This reaffirms the need to continue focusing on rapid initiation of bystander CPR and early defibrillation.

**Highlights:** *What is already known on this topic:*

- In coherent regions, key figures and trends of out-of-hospital cardiac arrest (OHCA) are vital to improve favourable outcomes.
- Up-to-date comprehensive OHCA data of the metropolitan area of Vienna, Austria, have been scarce.

*What this study adds:*

- This study provides an overview of OHCA key data between 2019 and 2023.
- General survival after OHCA in Vienna was stable but significant improvements in outcome parameters are seen in a ‘high outcome potential cohort’ over the last 15 years.

*How this study might affect research, practice or policy:*

- Stakeholders should continue to focus on rapid initiation of bystander CPR, early defibrillation, and high performance CPR by the emergency medical service.

## Background

The management and treatment of sudden out-of-hospital cardiac arrest (OHCA) remains a global health problem due to its critical time-dependent nature. It is a major challenge for emergency medical services (EMS) and, consequently, for emergency departments (ED) and intensive care units (ICU), continuing to depend on their infrastructure. The incidence rates of OHCA (or more exactly, the number of cases in which cardiopulmonary resuscitation (CPR) was performed), varies between 19 and 207 per 100,000 population per year in Europe. [1–3] Although survival rates have improved significantly over the years, survival to discharge remains low. Survival rates (survival to hospital discharge or survival at 30 days) for OHCA are reported to be between 8.0% and 11.3%, but still vary widely in communities. [1–4] This variability may be due to different cultural approaches to cardiac arrest, the presence of EMS support in both rural and urban regions, or awareness among the public. [2] Also, regional healthcare systems may potentially differ in their level of care and experience with cardiac arrest patients.

The most recent European study to report overview key figures is EuReCA Two: In 2017, 8% of patients survived to hospital discharge. [2] Specifically for the city of Vienna, Nürnberger et al. showed in 2009 to 2010 a survival rate to hospital discharge of 11.3% and a favourable neurological outcome in 8.7%. [3] As more than ten years have passed since this last statement, the study at hand aimed to analyze incidences and outcome trends of OHCA in the metropolitan area of Vienna over the last five years.

## Methods

### Study Design, Patients and Data Acquisition

The metropolitan area of Vienna has a population of around two million (as of January 2024) [5] and approximately 415km^2^. It is served by the Vienna Ambulance Service, which has been described in detail elsewhere. [6,7]

This retrospective study included all patients who suffered OHCA in Vienna, Austria, and in whom any CPR was started between 01/01/2019 and 31/12/2023. Exclusion criteria consisted of pure in-hospital cardiac arrest and pure declarations of death without CPR ever started. Pediatric and (presumed) traumatic cardiac arrest were not statistically evaluated in this publication and will be assessed in detail in the future *(Figure 1)*.

**Figure 1:**
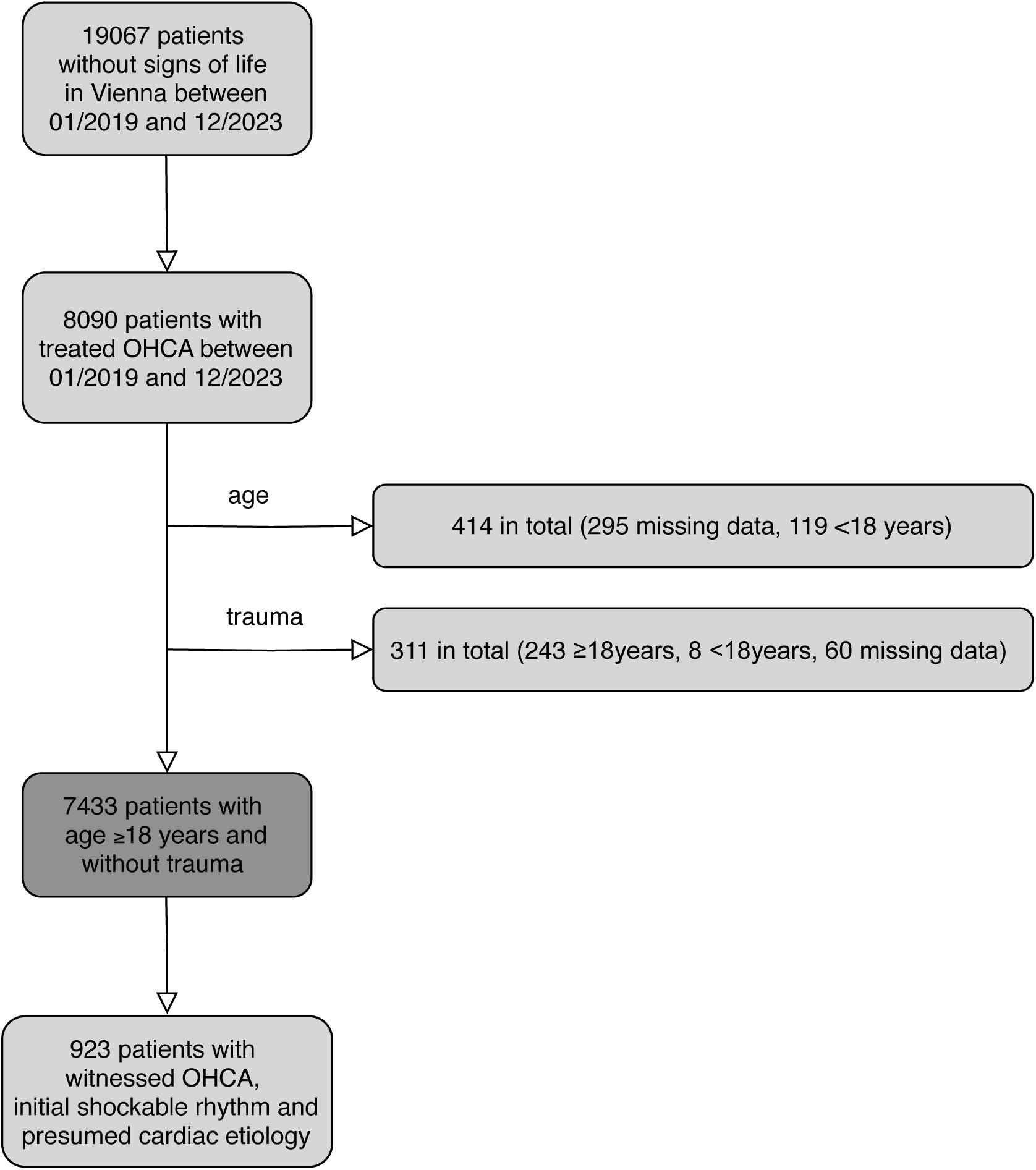
Patient flowchart. OHCA = out-of-hospital cardiac arrest.

Data were extracted from the pseudonymised standard ambulance run reports of the Emergency Medical Service Vienna, a physician-based professional EMS. All survivors of cardiac arrest were followed up to hospital discharge by assessing hospital records, discharge reports, and/or other clinical information. The outcome categories used were survival to hospital discharge and neurological outcome (favourable defined as a Cerebral Performance Category (CPC) 1 or 2, unfavourable 3 or 4 (death=5)) at hospital discharge. [8] Before the background of low subgroup patient numbers, CPC 1 and 2, as well as 3 and 4 were grouped to obtain more discriminatory power.

The study was approved by the Ethics Committee of the Medical University of Vienna (No. 1396/2020). It was conducted in accordance with the current version of the Declaration of Helsinki. The Ethics Committee waived the requirement for informed consent due to the retrospective nature of the study.

### COVID-19 dynamics

The coronavirus-disease 2019 (COVID-19) pandemic with respective governmental measures and lockdowns reached Austria in late February / early March 2020. Details and dynamics of epidemiological data concerning the pandemic within our observation period (starting with the first case on 25/02/2020 until the end of the last “hard” lockdown 03/05/2021) were obtained from official Austrian government records to assess the potential impact of COVID-19 on OHCA in Vienna. [9]

### Patient and Public Involvement

As this was a retrospective data analysis, patient and public involvement in the design was not applicable. Results will be dissmeminated via press statement and social media, as well as information on a public website.

### Statistical analysis

Variables are presented as absolute values (n), relative frequencies (%, with 95% confidence intervals (CI)) and medians with interquartile ranges (IQR). Comparisons between groups were made using the t-test, the Mann-Whitney U test, and the *χ*^2^- test. Population figures published by the Austrian / Viennese government were assessed to calculate incidences per year. [5] Poisson regression was used to analyze incidences before, during, and after the lockdowns in Vienna. Associations of survival and neurological outcome with potential explanatory factors were analyzed using simple logistic regression models. Odds ratios were calculated for each factor. Explanatory factors with odds ratios significantly different from 1 (at p≤0.05) were included in a multiple logistic regression model. The subgroup with the highest probability of survival (= patients with witnessed cardiac arrest of suspected cardiac etiology and an initial shockable rhythm [10]) was referred to as the ‘High outcome potential subgroup’. For data analysis, we used SPSS (IBM Corp. Released in 2020. IBM SPSS Statistics for Windows, version 27.0. Armonk, NY: IBM Corp). A two-tailed p-value of <0.05 was considered statistically significant.

## Results

At the beginning of 2024, Vienna had around 2 million inhabitants. [5] Between January 2019 and December 2023, 19067 (2019: 4210; 2020: 3795; 2021: 3661; 2022: 3864; 2023: 3627) patients without signs of circulation were assessed by the teams of the Vienna Emergency Medical Service. In 8090, cardiopulmonary resuscitation was initiated. After excluding traumatic and pediatric resuscitations (for details see *Figure 1*), a total of 7433 patients (77.1/100,000 inhabitants) were included in the main analysis *(Figure 1)*. This gives an annual incidence rate of 71.0/100,000 population for 2019, 78.0/100,000 for 2020, 78.0/100,000 for 2021, 83.0/100,000 for 2022 and 74.0/100,000 for 2023, with the highest incidence in the first political district of Vienna *(Supplementary Figure S1 and Supplementary Table S1)*. In the included study population (n=7433), the median age was 73 years (IQR 60-82) and patients were predominantly male (n=4449/7433; 59.9%). The most common suspected underlying cause of OHCA was a cardiac aetiology at 67.8% (n=5042/7433). Independently, hypoxia was suspected in 15.0% (n=1112/7433), pulmonary embolism in 6.8% (n=502/7433), non-traumatic haemorrhage in 3.4% (n=254/7433), intoxication in 2.3% (n=173/7433), a cerebral event in 1.3% (n=97/7433), drowning in 0.4% (n=33/7433), and other/unknown cause in 3% (n=220/7433). The initial rhythm established by the rescuers was shockable in 17.5% (n=1298/7433; ventricular fibrillation 14.7% (n=1096/7433)). Cardiac arrest was witnessed (by laypersons or medical professionals) in 52.5% (n=3904/7433), with 67.8% of bystanders being laypersons (n=2648/3906). In 33.7% of all cases (n=2506/7433), layperson bystander CPR was initiated. This represents 94.6% (n=2506/2648) of all layperson witnessed CPRs. OHCA occurred in private residences in 81.6% (n=6064/7433). Details are shown in *Table 1*. The median time from the onset of cardiac arrest / collapse to the arrival of the first ambulance on scene was nine minutes ((IQR 6-12); mean 10.1 minutes, SD 13.4) and the median time from receipt of the emergency call to the arrival of the first ambulance on scene was nine minutes ((IQR 7-12 minutes); mean 9.9 minutes; SD 8.2).

**Table 1.** Characteristics of the study population, OHCA events and Outcome - overall. OHCA = Out-of-hospital cardiac arrest, EMS = emergency medical services, CPR = cardiopulmonary resuscitation, ROSC = Return of spontaneous circulation, CPC = Cerebral Performance Categories, IQR= interquartile range, VF = Ventricular fibrillation, VT = Ventricular tachycardia, * Sustained ROSC = ROSC >20min.

|  |  | Overall<br>(n=7433) | 2019<br>(n=1354) | 2020<br>(n=1497) | 2021<br>(n=1499) | 2022<br>(n=1608) | 2023<br>(n=1475) | p-value |
| --- | --- | --- | --- | --- | --- | --- | --- | --- |
| Incidence | 100,000/<br>inhabitants/<br>year | 77.1 | 71.0 | 78.0 | 78.0 | 83.0 | 74.0 |  |
| Characteristics of the study population, OHCA events |  |  |  |  |  |  |  |  |
| Sex – female | n (%) | 2984 (40.1) | 540 (39.9) | 591 (39.5) | 613 (40.9) | 639 (39.7) | 601 (40.7) | 0.910 |
| Age – years | median (IQR) | 73.0 (22.0) | 73.0 (22.0) | 74.0 (23.0) | 73.0 (22.0) | 73.0 (21.0) | 74.0 (23.0) | 0.914 |
| BMI - kg/m<sup>2</sup> | median (IQR) | 26.1 (7.7) | 26.3 (7.4) | 26.2 (7.4) | 26.2 (7.8) | 26.0 (8.0) | 25.9 (6.5) | 0.009 |
| Location | n (%) |  |  |  |  |  |  |  |
| Home |  | 6064 (81.6) | 1081 | 1230 | 1253 | 1312 | 1188 | 0.086 |
| Public Place |  | 1321 (17.8) | (79.8) | (82.2) | (83.6) | (81.6) | (80.5) | 0.154 |
| Other |  | 48 (0.6) | 261 (19.3) | 257 (17.2) | 240 (16.0) | 286 (17.8) | 277 (18.8) |  |
|  |  |  | 12 (0.9) | 10 (0.7) | 6 (0.4) | 10 (0.6) | 10 (0.7) |  |
| EMS arrival time – minutes | median (IQR) | 9.0 (6.0) | 8.0 (5.0) | 9.0 (6.0) | 8.0 (6.0) | 9.0 (6.0) | 9.0 (6.0) | 0.611 |
| Witnessed | n (%) |  |  |  |  |  |  |  |
| No |  | 3527 (47.5) | 611 (45.1) | 693 (46.3) | 749 (50.0) | 778 (48.4) | 696 (47.2) | 0.086 |
| Yes, by bystander |  | 2648 (35.6) | 498 (36.8) | 524 (35.0) | 520 (34.7) | 554 (34.5) | 552 (37.4) | 0.324 |
| Yes, by EMS |  | 1256 (16.9) | 245 (18.1) | 280 (18.7) | 230 (15.3) | 275 (17.1) | 226 (15.3) | 0.039 |
| Unknown |  | 2 (0.0) | 0 | 0 | 0 | 1 (0.1) | 1 (0.1) |  |
| Layperson bystander CPR | n (%) | 2506 (33.7) | 440 (32.5) | 544 (36.3) | 482 (32.2) | 534 (33.2) | 506 (34.3) | 0.109 |
| Layperson AED use – turned on | n (%) | 123 (1.7) | 18 (1.3) | 20 (1.3) | 20 (1.3) | 33 (2.1) | 32 (2.2) | 0.151 |
| Layperson AED – shock delivered | n (%) | 52 (0.7) | 8 (0.6) | 12 (0.8) | 2 (0.1) | 18 (1.1) | 12 (0.8) | 0.020 |
| First rhythm |  |  |  |  |  |  |  |  |
| First shockable rhythm | n (%) | 1298 (17.5) | 277 (20.5) | 286 (19.1) | 233 (15.5) | 261 (16.2) | 241 (16.3) | 0.001 |
| VF |  | 1096 | 237 | 240 | 214 | 215 | 190 |  |
| VT |  | (14.7) | (17.5) | (16.0) | (14.3) | (13.4) | (12.9) |  |
| Unknown |  | 109 | 19 (1.4) | 21 (1.4) | 16 (1.1) | 23 (1.4) | 30 (2.0) | 0.001 |
| First non-shockable rhythm |  | (1.5) | 21 (1.6) | 25 (1.7) | 3 (0.2) | 23 (1.4) | 21 (1.4) |  |
|  |  | 93 (1.3) | 1075 | 1210 | 1264 | 1346 | 1230 |  |
| PEA |  | 6125 | (79.4) | (80.8) | (84.3) | (83.7) | (83.4) |  |
| Asystolie |  | (82.4) | 402 | 419 | 432 | 472 | 402 |  |
| Unknown |  | 2127 | (29.7) | (28.0) | (28.8) | (29.4) | (27.3) |  |
| Unknown |  | (28.6) | 620 | 744 | 782 | 831 | 778 |  |
|  |  | 3755 | (45.8) | (49.7) | (52.2) | (51.7) | (52.7) |  |
|  |  | (50.5) | 53 (3.9) | 47 (3.1) | 50 (3.3) | 43 (2.6) | 50 (3.4) |  |
|  |  | 243 | 2 (0.1) | 1 (0.1) | 2 (0.1) | 1 (0.1) | 4 (0.3) |  |
|  |  | (3.3) |  |  |  |  |  |  |
|  |  | 10 (0.1) |  |  |  |  |  |  |
| Suspected cardiac aetiology | n (%) | 5042 (67.8) | 935 (69.1) | 1062 (70.9) | 1016 (67.8) | 1049 (65.2) | 980 (66.4) | 0.008 |
| Time to (minutes) . . . |  |  |  |  |  |  |  |  |
| first chest compressions | median (IQR) | 3.0 (9.0) | 3.0 (9.0) | 3.0 (8.0) | 4.0 (9.0) | 3.0 (9.0) | 4.0 (9.0) | 0.005 |
| first defibrillation | median (IQR) | 10.0 (10.5) | 10.0 (12.0) | 10.0 (11.0) | 10 .0 (12.0) | 9.0 (10.0) | 11.0 (10.0) | 0.654 |
| first adrenaline | median (IQR) | 17.0 (10.0) | 16.0 (9.0) | 17.0 (11.0) | 17.0 (11.0) | 17.0 (11.0) | 17.0 (11.0) | 0.234 |
| Airway managment |  |  |  |  |  |  |  |  |
| Endotracheal intubation | n (%) | 4591 (61.8) | 810 (59.8) | 909 (60.7) | 938 (62.6) | 995 (61.9) | 939 (63.7) | 0.238 |
| Supraglottic device | n (%) | 561 (7.5) | 150 (11.1) | 121 (8.1) | 104 (6.9) | 95 (5.9) | 91 (6.2) | <0.001 |
| Outcome |  |  |  |  |  |  |  |  |
| Any ROSC | n (%) | 2403 (32.3) | 487 (36.0) | 464 (31.0) | 454 (30.3) | 521 (32.4) | 477 (32.3) | 0.015 |
| Sustained ROSC* | n (%) | 1840 (24.8) | 337 (27.8) | 366 (24.4) | 329 (21.9) | 390 (24.3) | 378 (25.6) | 0.007 |
| Survival to hospital discharge | n (%) | 689 (9.3) | 159 (11.7) | 142 (9.5) | 111 (7.4) | 138 (8.6) | 139 (9.4) | 0.002 |
| CPC 1/2 at hospital discharge | n (%) | 500 (6.8) | 112 (8.3) | 108 (7.2) | 81 (5.4) | 105 (6.5) | 94 (6.4) | 0.048 |
| CPC 3/4 at hospital discharge | n (%) | 129 (1.7) | 19 (1.4) | 24 (1.6) | 26 (1.7) | 23 (1.4) | 37 (2.5) | 0.048 |
| CPC missing at hospital discharge | n (%) | 60 (0.8) | 28 (2.0) | 10 (0.7) | 4 (0.3) | 10 (0.6) | 8 (0.5) |  |

### Outcome

In the main collective (n=7433), any ROSC was observed in 32.3% (n=2403/7433) of cases, while sustained ROSC was observed in 24.8% (n=1840/7433). 689 of 7433 patients (9.3%) were discharged from hospital alive, with 500 patients (6.8%) having a favourable neurological outcome (CPC 1 and 2). The detailed development over the years is shown in *Table 1 and Figure 2*. In the ‘High outcome potential subgroup’ (witnessed OHCA, suspected cardiac etiology and an initial shockable rhythm; n=923) any ROSC was seen in 67.5% (n=623/923; compared to the overall cohort p<0.001), sustained ROSC in 59.3% (n=547/923; p<0.001), survival to hospital discharge in 39.0% (n=360/923; p<0.001) and a favorable neurological outcome in 29.6% (n=273/923; p<0.001). The detailed development over the years for this subgroup is shown in *Table 2 and Figure 2*. When looking only at those patients who had a CPC of 1 or 2 (n=5836) prior to resuscitation (and therefore presumably only few impairments in daily life), a moderately better outcome can be seen than in the main collective (survival to hospital discharge 11.3%, favourable neurological outcome in 8.3% *(Supplementary Table S2)*. Between the first (18-64 years; n=2405) and third (≥80 years; n=2502) age tertiles of this patient cohort, survival to hospital discharge (17.0%, n=408/2405 vs. 2.7%, n=68/2502) and favourable neurological outcome (12.4%, n=299/2405 vs. 1.9%, n=48/2502) decrease significantly *(Supplementary Table S3)*.

**Figure 2:**
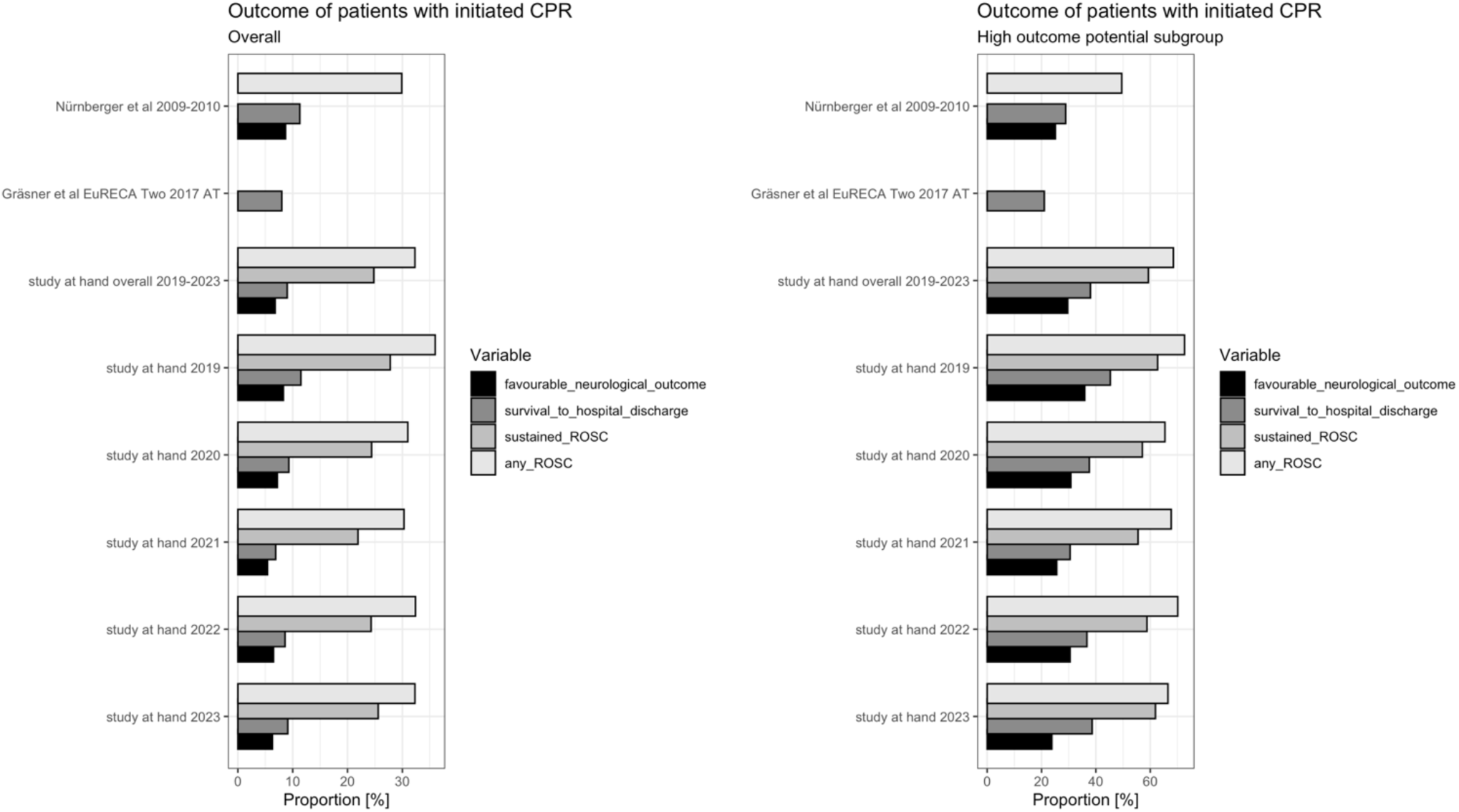
Outcome for patients (overall (n=7433) and high outcome potential subgroup (n=923) in whom cardiopulmonary resuscitation was initiated. AT = Austria, ROSC = Return of spontaneous circulation.

**Table 2.** Characteristics of the high outcome potential subgroup (witnessed, suspected cardiac aetiology and an initial shockable rhythm) - OHCA events and Outcome. OHCA = Out-of-hospital cardiac arrest, EMS = emergency medical services, CPR = cardiopulmonary resuscitation, ROSC = Return of spontaneous circulation, CPC = Cerebral Performance Categories, IQR= interquartile range, * Sustained ROSC = ROSC >20min.

|  |  | Overall<br>(n=923) | 2019<br>(n=201) | 2020<br>(n=205) | 2021<br>(n=164) | 2022<br>(n=177) | 2023<br>(n=176) | p-value |
| --- | --- | --- | --- | --- | --- | --- | --- | --- |
| Characteristics of the high outcome potential subgroup, OHCA events |  |  |  |  |  |  |  |  |
| Sex – female | n (%) | 202 (21.9) | 38 (18.9) | 47 (22.9) | 40 (24.4) | 38 (21.5) | 39 (22.2) | 0.772 |
| Age – years | median<br>(IQR) | 64.0 (20.0) | 64 (19.0) | 66.0 (22.0) | 63.0 (21.0) | 67.0 (22.0) | 62.5<br>(21.75) | 0.778 |
| BMI - kg/m<sup>2</sup> | median<br>(IQR) | 27.7 (6.5) | 27.8 (6.2) | 27.7 (6.2) | 26.6 (7.7) | 27.8 (8.4) | 27.7 (6.9) | 0.843 |
| Location | n (%) |  |  |  |  |  |  |  |
| Home |  | 499 (54.1) | 104 (51.7) | 113 (55.1) | 93 (56.7) | 101 (57.1) | 88 (50.0) | 0.592 |
| Public Place |  | 407 (44.1) | 90 (44.8) | 88 (42.9) | 70 (42.7) | 74 (41.8) | 85 (48.3) | 0.751 |
| Other |  | 17 (1.8) | 7 (3.5) | 4 (2.0) | 1 (0.6) | 2 (1.1) | 3 (1.7) |  |
| EMS arrival time – minutes | median<br>(IQR) | 8.0 (5.0) | 8.0 (4.5) | 8.0 (5.0) | 8.0 (5.0) | 8.0 (5.0) | 8.0 (4.5) | 0.319 |
| Witnessed | n (%) |  |  |  |  |  |  |  |
| Yes, by bystander |  | 732 (79.3) | 153 (76.1) | 163 (79.5) | 131 (79.9) | 141 (79.7) | 144 (81.8) | 0.741 |
| Yes, by EMS |  | 190 (20.6) | 48 (23.9) | 42 (20.5) | 33 (20.1) | 36 (20.3) | 31 (17.6) | 0.678 |
| Layperson bystander CPR | n (%) | 475 (51.5) | 91 (45.3) | 107 (52.2) | 85 (51.8) | 95 (53.7) | 97 (55.1) | 0.352 |
| Layperson AED use – turned on | n (%) | 48 (5.2) | 8 (4.0) | 11 (5.4) | 4 (2.4) | 12 (6.8) | 13 (7.4) | 0.218 |
| Layperson AED use – shock delivered | n (%) | 42 (4.6) | 8 (4.0) | 10 (4.9) | 2 (1.2) | 12 (6.8) | 10 (5.7) | 0.139 |
| Time to (minutes) . . . |  |  |  |  |  |  |  |  |
| first chest compressions | median<br>(IQR) | 3.0 (6.0) | 3.0 (7.0) | 3.0 (6.0) | 3.0 (6.0) | 4.0 (6.0) | 3.5 (7.0) | 0.877 |
| first defibrillation | median<br>(IQR) | 8.0 (7.0) | 8.0 (8.0) | 8.0 (9.0) | 8.0 (7.3) | 8.0 (6.5) | 9.0 (7.0) | 0.767 |
| first adrenaline | median<br>(IQR) | 17.0 (7.0) | 17.0 (7.0) | 17.0 (8.0) | 17.0 (8.0) | 18.0 (6.0) | 17.0 (7.0) | 0.560 |
| Airway managment |  |  |  |  |  |  |  |  |
| Endotracheal<br>intubation | n (%) | 748 (81.0) | 164 (81.6) | 163 (21.8) | 135 (82.3) | 138 (78.0) | 148 (84.1) | 0.613 |
| Supraglottic device | n (%) | 66 (7.2) | 19 (9.5) | 19 (9.3) | 8 (4.9) | 10 (5.6) | 10 (5.7) | 0.245 |
| Outcome |  |  |  |  |  |  |  |  |
| Any ROSC | n (%) | 623 (67.5) | 146 (72.6) | 134 (65.4) | 111 (67.7) | 124 (70.1) | 117 (66.5) | 0.540 |
| Sustained ROSC* | n (%) | 547 (59.3) | 126 (62.7) | 117 (57.1) | 91 (55.5) | 104 (58.8) | 109 (61.9) | 0.577 |
| Survival to hospital<br>discharge | n (%) | 360 (39.0) | 91 (45.3) | 79 (38.5) | 53 (32.3) | 66 (37.3) | 71 (40.3) | 0.148 |
| CPC 1/2 at hospital<br>discharge | n (%) | 273 (29.6) | 72 (35.8) | 63 (30.7) | 42 (25.6) | 54 (30.5) | 42 (23.9) | 0.930 |
| CPC 3/4 at hospital<br>discharge | n (%) | 53 (5.7) | 4 (2.0) | 11 (5.4) | 8 (4.9) | 8 (4.6) | 22 (12.5) | 0.930 |
| CPC missing at<br>hospital discharge | n (%) | 34 (3.7) | 15 (7.5) | 5 (2.4) | 3 (1.8) | 4 (2.2) | 7 (3.9) |  |

### Association between outcome and underlying factors

Increasing age and cardiac arrest at home decreased the likelihood of sustained ROSC, survival to hospital discharge, and favourable neurological outcome (CPC 1/2 vs 3/4/5) (Supplementary *Table S4*). On the other hand, a witnessed cardiac arrest, an initially shockable rhythm, endotracheal intubation, and a shorter time to first professional ventilation increase these outcome probabilities significantly (Supplementary *Table S4*). A suspected cardiac aetiology and a lower time to the administration of amiodarone increased all favourable outcome probabilities except sustained ROSC, and the use of a supraglottic device diminished all favourable outcome probabilities except sustained ROSC and CPC 1/2. The mentioned variables remained statistically significant regarding favourable neurological outcome as well as survival to hospital discharge in a multivariate analysis, with the exception of suspected cardiac cause.

However, if only those patients with CPC 3 and 4 (without CPC 5 which is brain death) are considered for the poor neurological outcome group, age and witnessed cardiac arrest no longer have a significant influence. In addition to initial shockable rhythm (OR 1.65, CI 1.11-2.43, p=0.013), location at home (OR 0.65, CI 0.44-0.97, p=0.035), suspected cardiac cause (OR 1.76, CI 1.16-2.66, p=0.008), and endotracheal intubation (OR 1.24, CI 0.13-0.44, p<0.001), this subgroup was significantly affected by the time to first professional ventilation (OR 0.93, CI 0.89-0.97, p<0.001) and the time to the first amiodarone administration (OR 0.94, CI 0.89- 0.99, p=0.012). The time to first chest compression and first defibrillation was longer in patients with unfavourable outcomes, but the respective differences did not reach statistical significance. Details are shown in *Supplementary Table S4*.

### The influence of COVID-19

During the COVID-19 pandemic, a decrease in survival to hospital discharge was observed from 11.4% (n=193/1699) before the first lockdown to 8.4% (n=141/1679) during the lockdowns (p=0.004). The same was observed for any ROSC (35.1%, n=597/1699 vs. 29.8%, n=501/1679; p=0.001), sustained ROSC (27.0%, n=459/1699 vs. 23.3%, n=391/1679; p=0.014), and favourable neurological outcome (7.9%, n=135/1699 vs. 6.6%, n=110/1679; p=0.120). After the last lockdown in December 2023, outcome parameters increased again, but did not reach pre-pandemic levels *(Supplementary Table S5)*. Poisson regression analysis for the months stratified in during and after national lockdown revealed a significant trend towards an increase in the incidence of cardiac arrest (rate ratio 8.31, CI 8.28 – 8.338, p<0.01).

## Discussion

The present data describe the incidence and outcome of OHCA in Vienna, the development over the last five years, and the impact of the COVID-19 pandemic.

The overall incidence of OHCA where any CPR was started was 77.1 per 100,000 population per year. In 2009 and 2010, Nürnberger et al. [3] reported an incidence of 42.6 per 100,000 inhabitants per year for patients with cardiac arrest treated by the Emergency Medical Service Vienna. Looking beyond Vienna’s numbers to Europe as a whole, the EuReCA One study in 2014 [1] showed an incidence of 49 per 100,000 population per year, and the EuReCA Two study in 2017 [2] reported an incidence of 56 per 100,000 population per year. On a global scale, these figures are similar to those of Beck et al. in Australia and New Zealand at 47.6 per 100,000 inhabitants per year in 2015. [11] The incidence in our study is therefore moderately higher than it was in Vienna 15 years ago and is also higher than in international comparison. Reasons for this increase could be that this patient population includes all patients for whom resuscitation was ever initiated, even if it was quickly discontinued (e.g., at EMS arrival after initial bystander CPR), or due to a successful “culture of layperson resuscitation” in Vienna resulting from long-standing awareness campaigns and public CPR education. [12]

Survival to hospital discharge was 9.3% in the overall study population, compared with 11.3% in 2009-2010 [3] and 10.8% in 1990 [13]. It was 10% in the EuReCA One study [1] and 8% in the EuReCA Two study [2]. The most recent outcome study on OHCA by Jerkeman et al. shows survival to hospital discharge in 10.1% of cases. [4] In the present study, any ROSC was observed in 32.3% of cases. These figures are similar to the results of the EuReCA Two study and Nürnberger et al (32.7% [2] and 29.9% [3]). Similar results were also seen for sustained ROSC (24.8% in 2019-2023 and 24.9% in 2009-2010 [3]). Related to survival to hospital discharge, slightly fewer patients in Vienna were discharged with a favourable neurological outcome between 2019 to 2023 and 2009 to 2010 [3] (6.8% vs. 8.7%). However, on average, patients in our study were five years older than those in Nürnberger et al. [3] and in the EuReCA Two study (73 vs 68 vs 67.6 years) [2]. It is therefore reasonable to assume that resuscitation attempts have been extended to (more) elderly individuals in recent years. As a result, we then focused our analysis on those patients who appear generally more likely to survive OHCA on the one-, and those who had had a presumed good quality of life before resuscitation (CPC 1/2) on the other hand. In patients with witnessed cardiac arrest, an initial shockable rhythm and a suspected cardiac cause, our outcomes were clearly better than in previous studies. Any ROSC was seen in 67.5%, sustained ROSC in 59.3%, survival to hospital discharge in 39.0% and a favorable neurological outcome in 29.6%. This represents an improvement over the last decade compared with both the EuReCA Two study (any ROSC 59%, survival to hospital discharge 31%) and Nürnberger et al. (any ROSC 49.5%, survival to hospital discharge 28.9%, CPC 1/2 25.1%). [2,3] Patients with a suspected CPC 1/2 before resuscitation had better outcomes than the overall cohort (11.3% survival to hospital discharge, 8.3% good neurological outcome), but not as good as the high-potential subgroup. The idea behind the formation of the subgroup of patients living with a CPC 1 or 2 before their OHCA was to exclude those patients who had a low chance of surviving the OHCA in the first place, due to extensive comorbidities, for example bed-ridden nursing home residents.

In the years between 2019 and 2023, there was a noticeable decrease in the outcome parameters. In 2019, 11.7% of patients were discharged from hospital after an OHCA, compared to 9.5% in 2020 and 7.4% in 2021. The same applied to a favourable neurological outcome: 8.3% of patients were discharged with a CPC 1/2 in 2019, 7.2% in 2020 and 5.4% in 2021. From 2022 onwards, these figures increase slightly, but did not reach the original values. The COVID-19 pandemic which reached Vienna in February 2020 is considered as a possible cause. The lockdowns reduced survival to discharge from hospital from 11.4% to 8.4% and favourable neurological outcome from 7.9% to 6.6%. This reduction has also been demonstrated by Ball et al (11.7% vs. 6.1%). [14] Whether this was due to excess mortality in patients with COVID-19 or other reasons is not fully understood. However, it has been suggested that during the pandemic, OHCAs occurred more frequently at home, emergency services took longer to arrive, fewer AEDs were used, and there were delays in key interventions like time to first defibrillations or time to first epinephrine. [14,15]

Similarly to the study results of Nürnberger et al. [3], we could demonstrate increasing age and cardiac arrest at home to be unfavourable for the outcome, while an witnessed cardiac arrest, an initial shockable rhythm, and a suspected cardiac cause were favourable influencing factors. This demonstrates once again how important it is to focus on increasing the rate of bystander-initiated CPR, the quality of layperson resuscitation, and early defibrillation. Also, our data showed a beneficial influence of early professional ventilation and endotracheal intubation, as well as early medication application, highlighting the importance of a well-functioning EMS. The Austrian Cardiac Arrest Awareness Association is promoting projects in Vienna in this regard through lay resuscitation courses and the expansion of first responder systems, e.g., by equipping the Vienna police with AEDs. [16,17] Future health policies could further support these endeavours, for instance a nation-wide CPR registry [18], mandatory CPR training for schoolchildren [19], or real BLS courses for obtaining a driving license [20]. Naturally, further research is vital for improving pre-hospital and in-hospital care, and should include co-morbidities, socioeconomic status, chronic medication, or details of post-ROSC care.

### Limitations

The main limitation of this analysis is that, in contrast to the Core Outcome Set for Cardiac Arrest (COSCA) [8], only survival and neurological function (CPC) were assessed, and health-related quality of life was not included due to organizational and limited resources. Another point is that although a decrease in survival was observed during the COVID-19 pandemic, it remains unknown whether these patients were COVID-19 positive. Moreover, we did not assess patient comorbidities or socioeconomical status as we did not have the respective data available. Another major limitation is that patient outcomes were only followed up until hospital discharge. Also, our results are only directly applicable to the city of Vienna. Data from other countries, regions, or event continents (such as reported, for example in the EuReCA studies [2]) will differ. Of note, the differences between our data and the reported Austrian data in the EuReCA studies stem from a different catchment area (only Vienna vs. several Austrian regions together).

## Conclusion

This study analysed the incidence and outcome of OHCA in the city of Vienna over the last five years. Although survival rates after OHCA continue to be lower than striven for, there have been notable improvements in outcomes over the past 15 years, especially for a high-potential patient cohort. This progress underscores the crucial importance of maintaining a strong emphasis on the immediate initiation of bystander CPR, early defibrillation, and high-performance CPR by the EMS, which are vital factors for enhancing patient survival and recovery. Respective health policies (e.g., a nation-wide CPR registry and mandatory CPR training for schoolchildren or for obtaining a driving licence) could further boost CPR research and favourable patient outcomes.

## Declarations

### Data availability

The study data are available from the corresponding author upon reasonable request in accordance to national law and organizational rules.

## Acknowledgements

The authors want to highlight the meticulous and tireless work of all healthcare personnel involved in cardiac arrest care in Vienna. Without this multiprofessional group, the presented outcomes would not be possible. In addition, the contributions to patient care, system organisation, and/or data assessment of the following individuals must be acknowledged: Rainer Gottwald, Rene Adler, Harald Bendl, Michael Bösze-Schaffer, Christoph Cincera, Sabine Dunkl, Benedikt Faulhammer, Mathias Gatterbauer, Wolfgang Klobouchnik, Philipp Gonzo, Jörg Holzinger, Carmen Huber, Harry Kopietz, Ronald Kopta, Michael Mareda, Henrik Maszar, Jürgen Novotny, Marcel Pfaffenlehner, Melanie Puhr, Julian Raming, Bernhard Saxinger, Bertram Schadler, Peter Scherer, Andreas Schindler, Alexander Schussmann, Martin Thalhammer, Günther Thiel, and Julia Zielinski.

## References

1. Gräsner JT, Lefering R, Koster RW, Masterson S, Böttiger BW, Herlitz J, u. a. EuReCa ONE-27 Nations, ONE Europe, ONE Registry: A prospective one month analysis of out-of-hospital cardiac arrest outcomes in 27 countries in Europe. Resuscitation. August 2016;105:188–95.

2. Gräsner JT, Wnent J, Herlitz J, Perkins GD, Lefering R, Tjelmeland I, u. a. Survival after out-of-hospital cardiac arrest in Europe - Results of the EuReCa TWO study. Resuscitation. 1. März 2020;148:218–26.

3. Nürnberger A, Sterz F, Malzer R, Warenits A, Girsa M, Stöckl M, u. a. Out of hospital cardiac arrest in Vienna: incidence and outcome. Resuscitation. Januar 2013;84(1):42–7.

4. Jerkeman M, Sultanian P, Lundgren P, Nielsen N, Helleryd E, Dworeck C, u. a. Trends in survival after cardiac arrest: a Swedish nationwide study over 30 years. Eur Heart J. 7. Dezember 2022;43(46):4817–29.

5. (https://www.wien.gv.at/statistik/bevoelkerung/bevoelkerungsstand/ retrieved on 22 July 2024 at 09:36).

6. https://www.wien.gv.at/gesundheit/einrichtungen/rettung/organisation/ retrieved on 6 August 2024 at 14:11.

7. Mueller M, Losert H, Sterz F, Gelbenegger G, Girsa M, Gatterbauer M, u. a. Prehospital emergency medicine research by additional teams on scene - Concepts and lessons learned. Resusc Plus. Dezember 2023;16:100494.

8. Haywood K, Whitehead L, Nadkarni VM, Achana F, Beesems S, Böttiger BW, u. a. COSCA (Core Outcome Set for Cardiac Arrest) in Adults: An Advisory Statement From the International Liaison Committee on Resuscitation. Circulation. 29. Mai 2018;137(22):e783–801.

9. https://www.data.gv.at/daten/covid-19/ Amtliches COVID-19 Dashboard des BMSGPK/AGES assessed 12.08.2024.

10. Thannhauser J, Nas J, Waalewijn RA, van Royen N, Bonnes JL, Brouwer MA, u. a. Towards individualised treatment of out-of-hospital cardiac arrest patients: an update on technical innovations in the prehospital chain of survival. Neth Heart J. Juli 2022;30(7–8):345–9.

11. Beck B, Bray J, Cameron P, Smith K, Walker T, Grantham H, u. a. Regional variation in the characteristics, incidence and outcomes of out-of-hospital cardiac arrest in Australia and New Zealand: Results from the Aus-ROC Epistry. Resuscitation. Mai 2018;126:49–57.

12. Schnaubelt S, Krammel M. PULS - Austrian Cardiac Arrest Awareness Association: An overview of a multi-tiered and multi-facetted regional initiative to save lives. Resusc Plus. September 2023;15:100453.

13. Gaul GB, Gruska M, Titscher G, Blazek G, Havelec L, Marktl W, u. a. Prediction of survival after out-of-hospital cardiac arrest: results of a community-based study in Vienna. Resuscitation. Oktober 1996;32(3):169– 76.

14. Ball J, Nehme Z, Bernard S, Stub D, Stephenson M, Smith K. Collateral damage: Hidden impact of the COVID-19 pandemic on the out-of-hospital cardiac arrest system-of-care. Resuscitation. November 2020;156:157–63.

15. Pennington B, Bell S, Wright A, Hill JE. Impact of COVID-19 on out-of-hospital cardiac arrest care processes. J Paramed Pract. 2. Februar 2023;15(2):74–7.

16. Krammel M, Lobmeyr E, Sulzgruber P, Winnisch M, Weidenauer D, Poppe M, u. a. The impact of a high-quality basic life support police-based first responder system on outcome after out-of-hospital cardiac arrest. PLoS One. 2020;15(6):e0233966.

17. Krammel M, Eichelter J, Gatterer C, Lobmeyr E, Neymayer M, Grassmann D, u. a. Differences in Automated External Defibrillator Types in Out-of-Hospital Cardiac Arrest Treated by Police First Responders. J Cardiovasc Dev Dis. 27. April 2023;10(5):196.

18. Graesner JT, Fu Wah Ho A, Dicker B. Resuscitation registries - Worldwide initiatives to deliver data for saving more life after cardiac arrest. Resuscitation Plus. 2024 Sep 30:20:100790. doi:10.1016/j.resplu.2024.100790.

19. Schroeder DC, Semeraro F, Greif R, Bray J, Morley P, Parr M, u.a. Kids save lives: Basic life support education for schoolchildren: A narrative review and scientific statement from the International Liaison Committee on Resuscitation. Circulation. 2023 Jun 13;147(24):1854–1868. doi:10.1161/CIR.0000000000001128.

20. Gamberini L, Schnaubelt S, Picardi M, Semeraro F, Monsieurs KG. Learn to drive, learn CPR: Advancing road safety and life-saving skills across Europe. Resuscitation. 2025 Jan 30:207:110526. doi:10.1016.j.resuscitation.2025.110526.

