## Supplemental Figure and Tables for "Incidence, outcome, and dynamics of out-of-hospital cardiac arrest in the city of Vienna between 2019 and 2023"

### Supplements

**Supplementary Figure S1:** Location of cardiac arrest - classification by Viennese districts (1-23) and the incidence (per 100,000 inhabitants) of cardiac arrest in Vienna in 2023, and hospitals receiving cardiac arrest patients in the observational period. *Modified with R, Original file 2021: CC BY 4.0 Statistik Austria data.statistik.gv.at (retrieved on 12/08/2024).*

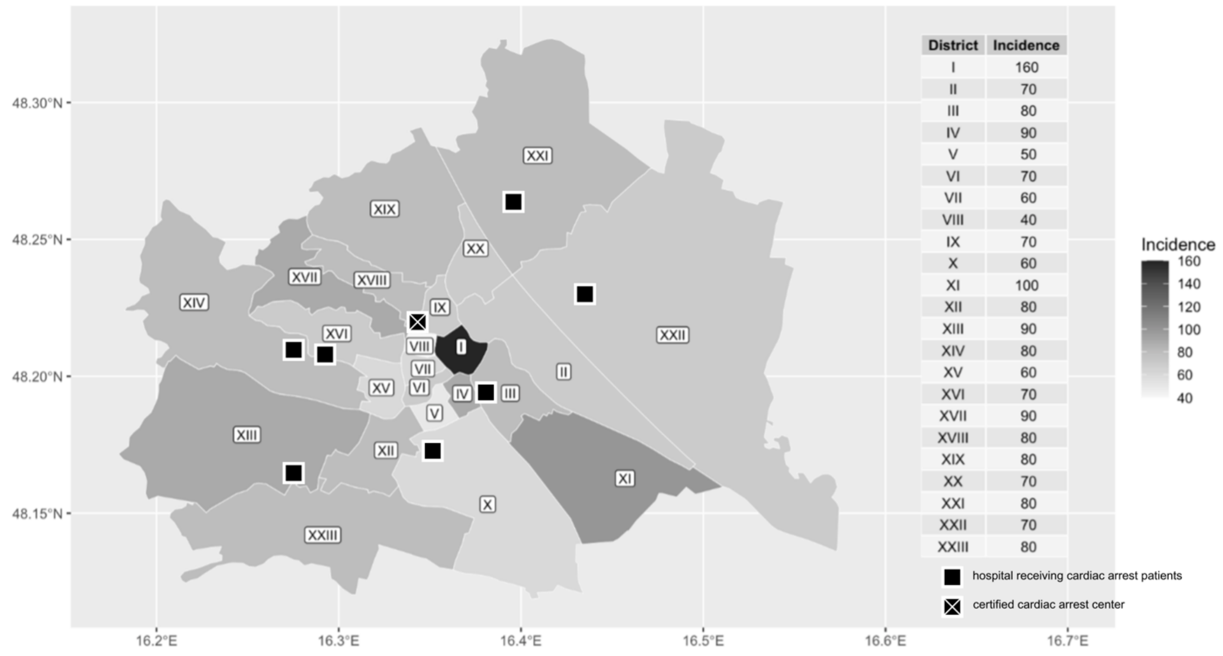

**Supplementary Table S1:** Location of cardiac arrest - classification by Viennese districts (1-23) and the incidence ([a]per 100,000 inhabitants) of cardiac arrest in Vienna over time

| **Districts** | | Overall (n=7433) | 2019 (n=1354) | 2020 (n=1497) | 2021 (n=1499) | 2022 (n=1608) | 2023 (n=1475) |
|---|---|---|---|---|---|---|---|
| 1st district | n (%) | 131 (1.8) | 31 (2.3) | 22 (1.5) | 21 (1.4) | 31 (1.9) | 26 (1.8) |
| 1st district | incidence[a] | | 190 | 140 | 130 | 200 | 160 |
| 2nd district | n (%) | 430 (5.8) | 82 (6.1) | 95 (6.3) | 84 (5.6) | 92 (5.7) | 77 (5.2) |
| 2nd district | incidence[a] | | 80 | 90 | 80 | 90 | 70 |
| 3rd district | n (%) | 379 (5.1) | 73 (5.4) | 82 (5.5) | 68 (4.5) | 74 (4.6) | 82 (5.6) |
| 3rd district | incidence[a] | | 80 | 90 | 70 | 80 | 80 |
| 4th district | n (%) | 133 (1.8) | 22 (1.6) | 24 (1.6) | 28 (1.9) | 29 (1.8) | 30 (2.0) |
| 4th district | incidence[a] | | 70 | 70 | 80 | 90 | 90 |
| 5th district | n (%) | 172 (2.3) | 29 (2.1) | 46 (3.1) | 33 (2.2) | 35 (2.2) | 29 (2.0) |
| 5th district | incidence[a] | | 50 | 80 | 60 | 70 | 50 |
| 6th district | n (%) | 118 (1.6) | 31 (2.3) | 23 (1.5) | 26 (1.7) | 17 (1.1) | 21 (1.4) |
| 6th district | incidence[a] | | 100 | 70 | 80 | 50 | 70 |
| 7th district | n (%) | 110 (1.5) | 20 (1.5) | 24 (1.6) | 20 (1.3) | 27 (1.7) | 19 (1.3) |
| 7th district | incidence[a] | | 60 | 80 | 60 | 90 | 60 |
| 8th district | n (%) | 68 (0.9) | 12 (0.9) | 12 (0.8) | 16 (1.1) | 19 (1.2) | 9 (0.6) |
| 8th district | incidence[a] | | 50 | 50 | 70 | 80 | 40 |
| 9th district | n (%) | 130 (1.7) | 23 (1.7) | 24 (1.6) | 30 (2.0) | 23 (1.4) | 30 (2.0) |
| 9th district | incidence[a] | | 50 | 60 | 70 | 60 | 70 |
| 10th district | n (%) | 768 (10.3) | 153 (11.3) | 161 (10.8) | 148 (9.9) | 172 (10.7) | 134 (9.1) |
| 10th district | incidence[a] | | 70 | 80 | 70 | 80 | 60 |

| | | | | | | | |
|---|---|---|---|---|---|---|---|
| 11th district | n (%) | 405 (5.4) | 74 (5.5) | 70 (4.7) | 86 (5.7) | 104 (6.5) | 71 (4.8) |
| 11th district | incidence[a] | | 70 | 70 | 80 | 100 | 100 |
| 12th district | n (%) | 405 (5.4) | 69 (5.1) | 78 (5.2) | 83 (5.5) | 94 (5.8) | 81 (5.5) |
| 12th district | incidence[a] | | 70 | 80 | 90 | 100 | 80 |
| 13th district | n (%) | 246 (3.3) | 41 (3.0) | 40 (2.7) | 52 (3.5) | 61 (3.8) | 52 (3.5) |
| 13th district | incidence[a] | | 80 | 70 | 100 | 110 | 90 |
| 14th district | n (%) | 382 (5.1) | 68 (5.0) | 71 (4.7) | 78 (5.2) | 88 (5.5) | 77 (5.2) |
| 14th district | incidence[a] | | 70 | 80 | 80 | 90 | 80 |
| 15th district | n (%) | 258 (3.5) | 44 (3.2) | 56 (3.7) | 57 (3.8) | 52 (3.2) | 49 (3.3) |
| 15th district | incidence[a] | | 60 | 70 | 70 | 70 | 60 |
| 16th district | n (%) | 356 (4.8) | 53 (3.9) | 72 (4.8) | 81 (5.4) | 75 (4.7) | 75 (5.1) |
| 16th district | incidence[a] | | 50 | 70 | 80 | 70 | 70 |
| 17th district | n (%) | 225 (3.0) | 32 (2.4) | 46 (3.1) | 39 (2.6) | 58 (3.6) | 50 (3.4) |
| 17th district | incidence[a] | | 60 | 80 | 70 | 100 | 90 |
| 18th district | n (%) | 175 (2.4) | 37 (2.7) | 32 (2.1) | 37 (2.5) | 29 (1.8) | 40 (2.7) |
| 18th district | incidence[a] | | 70 | 60 | 70 | 60 | 80 |
| 19th district | n (%) | 339 (4.6) | 61 (4.5) | 71 (4.7) | 58 (3.9) | 85 (5.3) | 64 (4.3) |
| 19th district | incidence[a] | | 80 | 100 | 80 | 120 | 80 |
| 20th district | n (%) | 319 (4.3) | 71 (5.2) | 51 (3.4) | 76 (5.1) | 59 (3.7) | 62 (4.2) |
| 20th district | incidence[a] | | 80 | 60 | 90 | 70 | 70 |
| 21th district | n (%) | 676 (9.1) | 121 (8.9) | 135 (9.0) | 141 (9.4) | 135 (8.4) | 144 (9.8) |
| 21th district | incidence[a] | | 70 | 80 | 80 | 80 | 80 |
| 22th district | n (%) | 750 (10.1) | 128 (9.5) | 156 (10.4) | 149 (9.9) | 159 (9.9) | 158 (10.7) |

| | | | | | | | |
|---|---|---|---|---|---|---|---|
| 22th district | incidence[a] | | 70 | 80 | 70 | 80 | 70 |
| 23th district | n (%) | 458 (6.2) | 79 (5.8) | 106 (7.1) | 88 (5.9) | 90 (5.6) | 95 (6.4) |
| 23th district | incidence[a] | | 70 | 100 | 80 | 80 | 80 |

**Supplementary Table S2:** Outcome – subgroup CPC 1 and 2 before cardiopulmonary resuscitation. ROSC = Return of spontaneous circulation, CPC = Cerebral Performance Categories, * Sustained ROSC = ROSC >20min

| **Outcome** | | Overall (n=5836) | 2019 (n=1131) | 2020 (n=1200) | 2021(n=1155) | 2022 (n=1228) | 2023 (n=1122) |
|---|---|---|---|---|---|---|---|
| Any ROSC | n (%) | 2156 (36.9) | 450 (39.8) | 426 (35.5) | 404 (35.0) | 447 (36.4) | 429 (38.2) |
| Sustained ROSC* | n (%) | 1689 (28.9) | 354 (31.3) | 344 (28.7) | 303 (26.2) | 343 (27.9) | 345 (30.7) |
| Survival to hospital discharge | n (%) | 660 (11.3) | 156 (13.8) | 138 (11.5) | 105 (9.1) | 127 (10.3) | 134 (11.9) |
| CPC 1/2 at hospital discharge | n (%) | 487 (8.3) | 111 (9.8) | 103 (8.6) | 80 (6.9) | 100 (8.1) | 93 (8.3) |
| CPC 3/4 at hospital discharge | n (%) | 113 (1.9) | 17 (1.5) | 24 (2.0) | 21 (1.8) | 17 (1.4) | 34 (3.0) |
| CPC missing at hospital discharge | n (%) | 60 (1.0) | 28 (2.5) | 11 (0.9) | 4 (0.3) | 10 (0.8) | 7 (0.6) |

**Supplementary Table S3:** Outcome categorised by age tertile. ROSC = Return of spontaneous circulation, CPC = Cerebral Performance Categories, * Sustained ROSC = ROSC >20min

| **Age 18-64** | | | | | | | |
|---|---|---|---|---|---|---|---|
| **Outcome** | | Overall (n=2405) | 2019 (n=432) | 2020 (n=498) | 2021 (n=487) | 2022 (n=502) | 2023 (n=486) |
| Any ROSC | n (%) | 978 (40.7) | 204 (47.2) | 199 (40.0) | 198 (40.7) | 189 (37.6) | 187 (38.5) |
| Sustained ROSC* | n (%) | 800 (33.3) | 168 (38.9) | 165 (33.1) | 157 (32.2) | 144 (28.7) | 166 (34.2) |
| Survival to hospital discharge | n (%) | 408 (17.0) | 92 (21.3) | 90 (18.1) | 70 (14.4) | 71 (14.1) | 85 (17.5) |
| CPC 1/2 at hospital discharge | n (%) | 299 (12.4) | 63 (14.6) | 67 (13.5) | 54 (11.1) | 57 (11.4) | 58 (11.9) |
| CPC 3/4 at hospital discharge | n (%) | 69 (2.9) | 12 (2.8) | 16 (3.2) | 13 (2.7) | 7 (1.4) | 21 (4.3) |
| CPC missing at hospital discharge | n (%) | 40 (1.7) | 17 (3.9) | 7 (1.4) | 3 (0.6) | 7 (1.4) | 6 (1.2) |
| **Age 65-79** | | | | | | | |
| **Outcome** | | Overall (n=2526) | 2019 (n=503) | 2020 (n=475) | 2021 (n=521) | 2022 (n=580) | 2023 (n=447) |
| Any ROSC | n (%) | 887 (35.1) | 194 (38.6) | 151 (31.8) | 159 (30.5) | 213 (36.7) | 170 (38.0) |
| Sustained ROSC* | n (%) | 670 (26.5) | 148 (29.4) | 121 (25.5) | 109 (20.9) | 161 (27.8) | 131 (29.3) |
| Survival to hospital discharge | n (%) | 213 (8.4) | 56 (11.1) | 39 (8.2) | 29 (5.6) | 51 (8.8) | 38 (8.5) |
| CPC 1/2 at hospital discharge | n (%) | 153 (6.1) | 41 (8.2) | 30 (6.3) | 18 (3.5) | 39 (6.7) | 25 (5.6) |
| CPC 3/4 at hospital discharge | n (%) | 45 (1.8) | 6 (1.2) | 8 (1.7) | 10 (1.9) | 9 (1.6) | 12 (2.7) |
| CPC missing at hospital discharge | n (%) | 15 (0.6) | 9 (1.8) | 1 (0.2) | 1 (0.2) | 3 (0.5) | 1 (0.2) |
| **Age >= 80** | | | | | | | |
| **Outcome** | | Overall (n=2502) | 2019 (n=419) | 2020 (n=524) | 2021 (n=491) | 2022 (n=526) | 2023 (n=542) |
| Any ROSC | n (%) | 539 (21.5) | 89 (21.2) | 114 (21.8) | 97 (19.8) | 119 (22.6) | 120 (22.1) |
| Sustained ROSC* | n (%) | 370 (14.6) | 61 (14.6) | 80 (15.3) | 63 (12.8) | 85 (16.2) | 81 (14.9) |
| Survival to hospital discharge | n (%) | 68 (2.7) | 11 (2.6) | 13 (2.5) | 12 (2.4) | 16 (3.0) | 16 (3.0) |

| | | | | | | | |
|---|---|---|---|---|---|---|---|
| CPC 1/2 at hospital discharge | n (%) | 48 (1.9) | 8 (1.9) | 11 (2.1) | 9 (1.8) | 9 (1.7) | 11 (2.0) |
| CPC 3/4 at hospital discharge | n (%) | 15 (0.6) | 1 (0.2) | 0 (0.0) | 3 (0.6) | 7 (1.3) | 4 (0.7) |
| CPC missing at hospital discharge | n (%) | 5 (0.2) | 2 (0.5) | 2 (0.4) | 0 (0.0) | 0 (0.0) | 1 (0.2) |

**Supplementary Table S4:** Univariate logistic regression for overall sustained ROSC, survival to hospital discharge and favourable neurological outcome. ROSC = Return of spontaneous circulation, OR = odds Ratio, CI = confidence interval, IQR= interquartile range, BMI = body mass index, CPC = Cerebral Performance Categories, LUCAS = mechanical chest compression system

| **Outcome – substained ROSC** | | | | | | |
|---|---|---|---|---|---|---|
| | | yes (n=1840) | no (n=5593) | p-value | OR | CI 95% |
| age (years) | median (IQR) | 68.0 (21.0) | 75 (22.0) | **<0.001** | 0.98 | 0.98 – 0.985 |
| BMI (kg/m$^2$) | median (IQR) | 27.3 (7.0) | 25.95 (8.0) | 0.306 | 1.00 | 0.99 – 1.00 |
| bystander witnessed | n (%) | 1418 (77.1) | 2488 (44.5) | **<0.001** | 4.03 | 3.59 – 4.51 |
| Shockable rhythm | n (%) | 696 (37.8) | 602 (10.8) | **<0.001** | 4.73 | 4.19 – 5.35 |
| location – home | n (%) | 1231 (66.9) | 4833 (86.4) | **<0.001** | 0.36 | 0.32 – 0.39 |
| suspected cardiac cause | n (%) | 1140 (62.0) | 3902 (69.8) | 0.061 | 0.76 | 0.68 – 1.02 |
| Endotracheal intubation | n (%) | 1572 (85.4) | 268 (14.6) | **<0.001** | 5.00 | 4.35 – 5.75 |
| Supraglottic device | n (%) | 120 (6.5) | 1720 (93.5) | 0.055 | 0.82 | 0.66 – 1.01 |
| **Time to (minutes) . . .** | | | | | | |
| first chest compressions | median (IQR) | 2.0 (7.0) | 5.0 (9.0) | 0.615 | 1.00 | 0.99 – 1.00 |
| first defibrillation | median (IQR) | 8.0 (9.0) | 13.0 (12.0) | 0.770 | 1.00 | 1.00 – 1.00 |
| first adrenaline | median (IQR) | 16.0 (10.0) | 17.0 (10.0) | 0.335 | 1.00 | 0.99 – 1.00 |
| first amiodarone | median (IQR) | 19.0 (10.0) | 23.0 (10.0) | 0.952 | 0.95 | 0.93 – 0.97 |
| first ventilation | median (IQR) | 9.0 (10.0) | 11.0 (12.0) | **<0.001** | 0.97 | 0.96 – 0.98 |
| first successful endotracheal intubation | median (IQR) | 18.0 (11.0) | 19.0 (11.0) | 0.065 | 0.99 | 0.99 – 1.00 |
| first successful supraglottic airway device | median (IQR) | 14.0 (12.0) | 16.0 (13.0) | 0.664 | 1.00 | 0.99 – 1.01 |
| start LUCAS | median (IQR) | 21.0 (15.3) | 24.0 (16.0) | 0.391 | 1.00 | 0.99 – 1.01 |
| switching on the defibrillator | median (IQR) | 6.0 (10.0) | 8.0 (12.0) | 0.075 | 1.00 | 0.99 – 1.00 |

| | | | | | | |
|---|---|---|---|---|---|---|
| first rhythm analysis | median (IQR) | 6.0 (10.0) | 9.0 (11.0) | 0.015 | 0.99 | 0.98 – 0.99 |
| **Outcome – hospital to survival** | | | | | | |
| | | yes (n=689) | no (n=6744) | p-value | OR | CI 95% |
| age (years) | median (IQR) | 61 (20) | 75 (21.0) | **<0.001** | 0.96 | 0.96 – 0.97 |
| BMI (kg/m$^2$) | median (IQR) | 27.3 (6.6) | 26.1 (8.0) | 0.834 | 1.00 | 1.00 – 1.00 |
| bystander witnessed | n (%) | 585 (84.9) | 3321 (49.2) | **<0.001** | 5.798 | 4.68 – 7.18 |
| Shockable rhythm | n (%) | 431 (62.6) | 867 (12.9) | **<0.001** | 11.34 | 9.55 – 13.43 |
| location – home | n (%) | 361 (52.4) | 5703 (84.6) | **<0.001** | 0.20 | 0.17 – 0.24 |
| suspected cardiac cause | n (%) | 513 (74.5) | 4529 (67.2) | **<0.001** | 1.42 | 1.19 – 1.70 |
| Endotracheal intubation | n (%) | 504 (73.1) | 185 (26.9) | **<0.001** | 1.77 | 1.49 – 2.11 |
| Supraglottic device | n (%) | 26 (3.8) | 663 (96.2) | **<0.001** | 0.46 | 0.31 – 0.68 |
| **Time to (minutes) . . .** | | | | | | |
| first chest compressions | median (IQR) | 1.0 (5.0) | 4.0 (10.0) | 0.391 | 0.99 | 0.99 – 1.00 |
| first defibrillation | median (IQR) | 6.0 (8.0) | 12.0 (12.0) | 0.429 | 1.00 | 1.00 – 1.00 |
| first adrenaline | median (IQR) | 15 (10.0) | 17.0 (10.0) | 0.302 | 0.99 | 0.99 – 1.00 |
| first amiodarone | median (IQR) | 18.0 (10.0) | 23.0 (11.0) | **<0.001** | 0.94 | 0.92 – 0.97 |
| first ventilation | median (IQR) | 7.0 (10.0) | 11.0 (11.0) | **<0.001** | 0.95 | 0.94 – 0.97 |
| first successful endotracheal intubation | median (IQR) | 18.0 (11.0) | 18.0 (11.0) | 0.731 | 0.99 | 0.99 – 1.01 |
| first successful supraglottic airway device | median (IQR) | 13.0 (10.5) | 16.0 (13.0) | 0.886 | 1.00 | 0.99 – 1.01 |
| start LUCAS | median (IQR) | 23.0 (15.3) | 23.0 (16.5) | 0.909 | 1.00 | 0.99 – 1.00 |
| switching on the defibrillator | median (IQR) | 4.0 (8.3) | 8.0 (11.0) | 0.831 | 1.00 | 1.00 – 1.00 |
| first rhythm analysis | median (IQR) | 5.0 (9.0) | 8.0 (11.0) | 0.842 | 1.00 | 1.00 – 1.00 |
| **Outcome - favourable cerebral outcome (CPC 1 or 2 vs CPC 3 or 4 or 5)** | | | | | | |

| | | yes (n=500) | no (n=6933) | p-value | OR | CI 95% |
|---|---|---|---|---|---|---|
| age (years) | median (IQR) | 61.0 (20.0) | 74.0 (22.0) | **<0.001** | 0.96 | 0.96 – 0.97 |
| BMI (kg/m$^2$) | median (IQR) | 27.3 (6.3) | 26.1 (8.0) | 0.860 | 1.00 | 1.00 – 1.00 |
| bystander witnessed | n (%) | 432 (86.4) | 3474 (50.1) | **<0.001** | 6.34 | 4.88 – 8.20 |
| Shockable rhythm | n (%) | 320 (64.0) | 978 (14.1) | **<0.001** | 10.83 | 8.91 – 13.15 |
| location – home | n (%) | 258 (51.6) | 5806 (83.7) | **<0.001** | 0.21 | 0.17 – 0.25 |
| suspected cardiac cause | n (%) | 380 (76.0) | 4662 (67.2) | **<0.001** | 1.54 | 1.25 – 1.91 |
| Endotracheal intubation | n (%) | 341 (68.2) | 159 (31.8) | **0.002** | 1.35 | 1.12 – 1.64 |
| Supraglottic device | n (%) | 16 (3.2) | 484 (96.8) | **<0.001** | 0.387 | 0.23 – 0.64 |
| **Time to (minutes) . . .** | | | | | | |
| first chest compressions | median (IQR) | 1.0 (4.0) | 4.0 (10.0) | 0.579 | 0.99 | 0.99 – 1.00 |
| first defibrillation | median (IQR) | 6.0 (8.0) | 11.0 (11.0) | 0.291 | 1.00 | 1.00 – 1.00 |
| first adrenaline | median (IQR) | 14.0 (9.0) | 17.0 (10.0) | 0.308 | 0.99 | 0.99 – 1.00 |
| first amiodarone | median (IQR) | 17.0 (8.75) | 22.0 (10.8) | **<0.001** | 0.92 | 0.89 – 0.95 |
| first ventilation | median (IQR) | 6.0 (10.0) | 11.0 (11.0) | **<0.001** | 0.95 | 0.93 – 0.96 |
| first successful endotracheal intubation | median (IQR) | 18.0 (11.0) | 18.0 (11.0) | 0.558 | 0.99 | 0.99 – 1.00 |
| first successful supraglottic airway device | median (IQR) | 13.0 (7.0) | 16.0 (13.0) | 0.910 | 1.00 | 0.99 – 1.01 |
| start LUCAS | median (IQR) | 21.0 (16.0) | 23.0 (16.0) | 0.899 | 1.00 | 0.99 – 1.01 |
| switching on the defibrillator | median (IQR) | 3.0 (8.0) | 8.0 (11.0) | 0.891 | 1.00 | 1.00 – 1.00 |
| first rhythm analysis | median (IQR) | 4.0 (8.0) | 8.0 (11.0) | 0.901 | 1.00 | 1.00 – 1.00 |
| **Outcome – favourable cerebral outcome (CPC 1 or 2 vs CPC 3 or 4)** | | | | | | |
| | | yes (n=500) | no (n=129) | p-value | OR | CI 95% |
| age (years) | median (IQR) | 62.0 (11.5) | 62.0 (11.5) | 0.219 | 0.99 | 0.98 – 1.01 |

| | | | | | | |
|---|---|---|---|---|---|---|
| BMI ($kg/m^2$) | median (IQR) | 27.3 (7.0) | 27.3 (7.0) | 0.671 | 0.99 | 0.99-1.01 |
| bystander witnessed | n (%) | 107 (82.9) | 107 (82.9) | 0.319 | 1.31 | 0.77 – 2.21 |
| Shockable rhythm | n (%) | 67 (51.9) | 67 (51.9) | **0.013** | 1.65 | 1.11 – 2.43 |
| location – home | n (%) | 80 (62.0) | 80 (62.0) | **0.035** | 0.65 | 0.44 – 0.97 |
| suspected cardiac cause | n (%) | 83 (64.3) | 83 (64.3) | **0.008** | 1.76 | 1.16 – 2.66 |
| Endotracheal intubation | n (%) | 341 (68.2) | 159 (31.8) | **<0.001** | 1.24 | 0.13 – 0.44 |
| Supraglottic device | n (%) | 16 (3.2) | 484 (96.8) | 0.119 | 0.50 | 0.21 – 1.20 |
| **Time to (minutes) . . .** | | | | | | |
| first chest compressions | median (IQR) | 1.0 (4.0) | 2.0 (6.0) | 0.790 | 1.00 | 0.99– 1.00 |
| first defibrillation | median (IQR) | 6.0 (8.0) | 9.0 (7.0) | 0.731 | 1.00 | 0.99 – 1.00 |
| first adrenaline | median (IQR) | 14.0 (9.0) | 17.0 (9.0) | 0.079 | 0.97 | 0.94 – 1.00 |
| first amiodarone | median (IQR) | 17.0 (8.8) | 22.0 (9.3) | **0.012** | 0.94 | 0.89 – 0.99 |
| first ventilation | median (IQR) | 6.0 (10.0) | 9.5 (7.8) | **<0.001** | 0.93 | 0.89 – 0.97 |
| first successful endotracheal intubation | median (IQR) | 18.0 (11.0) | 18.0 (11.0) | 0.951 | 0.99 | 0.98 – 1.02 |
| first successful supraglottic airway device | median (IQR) | 13.0 (7.0) | 10.0 (7.5) | 0.568 | 1.03 | 0.93 – 1.15 |
| start LUCAS | median (IQR) | 21.0 (16.0) | 25.0 (13.0) | 0.947 | 1.00 | 0.97 – 1.03 |
| switching on the defibrillator | median (IQR) | 3.0 (8.0) | 7.0 (9.0) | <0.001 | 0.93 | 0.90 – 0.97 |
| first rhythm analysis | median (IQR) | 4.0 (8.0) | 7.5 (8.3) | <0.001 | 0.94 | 0.90 – 0.97 |

**Supplementary Table S5:** The impact of COVID-19 on the incidence and outcome of OHCA in Vienna between 2019 and 2023. ROSC = Return of spontaneous circulation, CPC = Cerebral Performance Categories, * Sustained ROSC = ROSC >20min

| | | **Before the first lockdown (January 2019 until mid-March 2020)** | **During the first and last lowdown (Mid-March 2020 until May 2021)** | **After the last lockdown (May 2021 until December 2023)** |
|---|---|---|---|---|
| | | Overall (n=1699) | Overall (n=1679) | Overall (n= 4055) |
| age (years) | median (IQR) | 74.0 (23.0) | 73.0 (22.0) | 74.0 (12.0) |
| sex – female | n (%) | 690 (40.6) | 655 (39.0) | 1639 (40.4) |
| bystander witnessed | n (%) | 919 (54.1) | 892 (53.1) | 2095 (51.7) |
| Shockable rhythm | n (%) | 336 (19.8) | 299 (17.8) | 663 (16.4) |
| location – home | n (%) | 1366 (80.4) | 1396 (83.1) | 3302 (81.4) |
| suspected cardiac cause | n (%) | 1187 (69.9) | 1166 (69.4) | 2689 (66.3) |
| Time from cardiac arrest to arrival of the first ambulance at the scene (min) | median (IQR) | 8.0 (5.0) | 9.0 (7.0) | 9.0 (6.0) |
| Time to emergency call to arrival of the first ambulance at the scene (min) | median (IQR) | 9.0 (4.0) | 9.0 (5.0) | 9.0 (5.0) |
| **Outcome** | | | | |
| Any ROSC | n (%) | 597 (35.1) | 501 (29.8) | 1305 (32.2) |
| Sustained ROSC* | n (%) | 459 (27.0) | 391 (23.3) | 990 (24.4) |
| Survival to hospital discharge | n (%) | 193 (11.4) | 141 (8.4) | 355 (8.8) |
| CPC 1/2 at hospital discharge | n (%) | 135 (7.9) | 110 (6.6) | 255 (6.3) |
| CPC 3/4 at hospital discharge | n (%) | 21 (1.2) | 29 (1.7) | 79 (1.9) |
| CPC missing at hospital discharge | n (%) | 37 (2.2) | 2 (0.1) | 21 (0.5) |
